# A Multiple Linear Regression Analysis of Rural-Urban COVID-19 Risk Disparities in Texas

**DOI:** 10.1101/2021.01.05.20248921

**Authors:** Amber K. Luo, Sophia Zhong, Charles Sun, Jasmine Wang, Alexander White

## Abstract

As the number of COVID-19 cases in the U.S. rises, the differential impact of the pandemic in urban and rural regions becomes more pronounced, and the major factors relating to this difference remain unclear. Using the 254 counties of Texas as units of analysis, we utilized multiple linear regression to investigate the influence of 83 county-level predictor variables including race demographics, age demographics, healthcare and financial status, and prevalence of and mortality rate from COVID-19 risk factors on the incidence rate and case fatality rate from COVID-19 in Texas on September 15, 2020. Here, we report that urban counties experience, on average, 41.1% higher incidence rates from COVID-19 than rural counties and 34.7% lower case fatality rates. Through comparisons between our models, we found that this difference was largely attributable to four major predictor variables: namely, the proportion of elderly residents, African American residents, and Hispanic residents, and the presence of large nursing homes. According to our models, counties with high incidence rates of COVID-19 are predicted to have high proportions of African American residents and Hispanic residents coupled with low proportions of elderly residents. Furthermore, we found that counties with the highest case fatality rates are predicted to have high proportions of elderly residents, obese residents, and Hispanic residents, coupled with low proportions of residents ages 20-39 and residents who report smoking cigarettes. In our study, major variables and their effects on COVID-19 risk are quantified, highlighting the most vulnerable populations and regions of Texas.

## Introduction

The COVID-19 (Coronavirus Disease 2019) pandemic continues to exert major effects on healthcare systems and on the well-being of residents in both urban and rural regions. Texas is one of the hardest hit states by COVID-19, experiencing alarming surges in confirmed cases, hospitalizations, and deaths starting in mid June of 2020. As of September 15, 2020, there have been 15,785 deaths and 668,746 confirmed cases of COVID-19 across the 254 counties in Texas [1].

Previous studies have shown that urban counties in Texas experience significantly higher incidence rates and lower case fatality rates from COVID-19 when compared to rural counties [2]. Rural communities experience lower testing rates and higher positive test rates for COVID-19 [3], along with fewer financial resources [4], limited access to healthcare [5], and higher all-cause mortality rates in comparison to urban communities [6]. Furthermore, many hospitals in rural communities lack the ICU beds and primary care physicians necessary to deal with a local COVID-19 outbreak [7]. Rural counties also have higher prevalences of CDC (Centers for Disease Control and Prevention)-defined COVID-19 risk factors [8], including cardiovascular disease, chronic respiratory disease, obesity, and smoking, as well as higher rates of poverty and lower levels of physical activity [9]. Of those hospitalized with COVID-19, 75% have some underlying medical condition, regardless of age, such as diabetes, chronic respiratory diseases, and cardiovascular diseases [10]. Furthermore, rural and urban counties have many demographic differences, including a higher proportion of elderly and black residents in rural counties, populations that are thought to be especially vulnerable to COVID-19 [11]. These health and demographic disparities, among others, hold the key to understanding the differential impact of COVID-19 on urban and rural counties. Identifying the variables responsible for these differences and quantifying their effects on measures of COVID-19 risk will allow policy makers and healthcare professionals to determine the most suitable plans for COVID-19 based on the predicted vulnerability of a region.

Currently, few studies have investigated the relation of demographic differences in rural and urban counties to COVID-19 risk. Our study is the first to use multiple linear regression to definitively quantify the relationship of population characteristics to urban-rural COVID-19 disparities by creating models that explain the effect of various characteristics (predictor variables) on a single measure of COVID-19 risk (response variable). These multiple linear regression models were run for on two response variables: incidence rate from COVID-19 and case fatality rate from COVID-19. The county-level predictor variables we analyzed include, but are not limited to, prevalence of and mortality rate from CDC-defined COVID-19 risk factors, age demographics, race demographics, financial and living conditions, political leaning, and potential super-spreading sites. Our study highlights the most significant county-level variables in predicting COVID-19 incidence rate and case fatality rate and identifies the major underlying variables responsible for urban-rural disparities in COVID-19 risk.

## Data and Methods

### Data Sources

The units of analysis for this study are N = 254 counties in Texas. The COVID-19 Data Hub 2.2.0, accessed through the COVID19 Package in R, provided cumulative county-level reports of COVID-19 confirmed cases and deaths. Data was filtered to a 7 day window around September 15 (9/12 - 9/19), and the 7 day rolling averages of COVID-19 incidence rate and case fatality rate were calculated from this window. The 83 predictor variables and their sources can be grouped into the following four categories:

1. Population demographics: County-level data on percent population by five-year age strata from 0-85 years of age, race, education status, health insurance status, sex, unemployment status, and poverty status, as well as data on median income, population density per square mile, and the percentage of healthcare workers per county were obtained from the 2018 American Community Survey and the 2020 County Health Rankings [12, 13].
2. Health-related risk factors: County-level data on the prevalence of diabetes, cigarette smoking, obesity, and hypertension, as well as the mortality rate per 100,000 population from chronic respiratory disease, cardiovascular disease, alcohol use disorders, and drug use disorders were obtained from the IHME (Institute for Health Metrics and Evaluation) GHDx (Global Health Data Exchange) US Health Datasets. Data on survey responses on mask use, where participants from all counties in the US were asked to self-report how often they wear masks in public from 7/2/2020 - 7/14/2020, was obtained from the New York Times [14, 15].
3. Potential superspreading sites: Three discrete variables were created to examine the true effect of sites generally thought to have a higher incidence of COVID-19 on COVID-19 risk. These variables represented the presence of one or more meatpacking facilities, the presence of one or more prisons, and the presence of one or more large nursing homes (population *>*100) by county from Niche Meat Processing, Texas Almanac, and Texas Health and Human Services, respectively [16, 17, 18].
4. Political leaning: Due to the heavily politicized climate surrounding the COVID-19 pandemic, several measures of political leaning were collected. Data on the percentage of voters for Trump/Clinton in the 2016 presidential election and the percentage of voters for O’Rourke/Cruz in the 2018 Texas Senate election was obtained from the New York Times to estimate the true political leanings of the counties in Texas [19, 20].

All statistical analyses, models, and mapping of data were done using R version 1.3.959. A complete list of the predictor and response variables analyzed and their sources and dates can be found in Table 1.

### Statistical Analysis

Prior to model creation, counties were grouped into quintiles (1-5) by measures of risk: COVID-19 incidence rate per 100,000 population and case fatality rate, both calculated from 7-day rolling averages. Medians and IQRs were calculated for select predictor variables by quintile, and their significance in relation to COVID-19 risk was determined using Kruskal-Wallis tests, with a *p*-value of *<* 0.05 considered significant. These tables of COVID-19 risk quintiles (Table 2, 4) can be compared with a study by Khose, Moore, and Wang [2] using data from April 8, 2020.

### Models

Multiple Linear Regression (MLR) was used to model the effect of different subsets of predictor variables on each of the response variables. To investigate the difference between urban and rural counties, all models include the *Urban* variable, which denotes the urban-rural status of the county. 

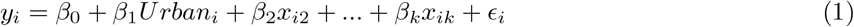

*y*_*i*_ represents either incidence rate or case fatality rate from COVID-19 for the *i*th county, and *x*_*ij*_ represents the value of the *j*th predictor variable for the *i*th county. The errors, *ϵ*_*i*_ ∼ *N* (0, *σ*^2^), are assumed independent.

The best subset of predictors for each response variable was chosen in two phases. In phase one, using the `LEAPS` package in R, all possible subsets of our 83 predictor variables were compared using the Bayesian information criteria. The optimal Box-Cox transformation was then fit to the best four models. In each case, the 95% confidence interval for the optimal power included 0, which suggested the natural log transformation. In phase two, the `LEAPS` package in R was used once more with the transformed response variable. The best four models for each response were then compared for strength of prediction, as determined by adjusted *R*^2^ and *F*-statistic; validity of prediction, as determined by normally distributed residual plots; and significance, as determined by the *p*-values of the variables included in the model and ANOVA tables. To assess possible multicollinearity, VIF (variance of inflation) factors were calculated for all predictors in our MLR models using the `car` package in R. In each case, the VIF never exceeded 2.744, indicating a lack of variable redundancy (threshold: VIF= 5).

## Results

### Incidence Rate Analysis

To elucidate the basic nature of the predictor variables in relation to COVID-19 risk and to provide for a deeper understanding of the coefficients in our MLR models, we analyzed the medians and IQRs of certain predictor variables by COVID-19 risk quintiles. Table 2 presents the distribution of county-level demographics and health outcomes by quintiles of COVID-19 incidence rates in Texas on September 15, 2020. Counties with higher incidence rates had generally higher median percentages of residents with diabetes (*p*-value: *<* .001), African American residents (*p*-value: .002), Hispanic residents (*p*-value: *<* .001), obese residents (*p*-value: *<* .001), residents 20-39 years of age (*p*-value: *<* .001), unemployed residents (*p*-value: .078) and generally higher population density per square mile (*p*-value: *<* .001). These counties also had generally lower median percentages of Caucasian American residents (*p*-value: .002) and elderly residents (*p*-value: *<* .001, *>* 70 years of age). In particular, the fifth quintile of COVID-19 incidence rate (*>* 2816.3 per 100,000 population) had the highest median percentage of residents beneath the poverty line (median: 16.6%; *p*-value: .002) and the lowest percentage of residents with health insurance (median: 97.7%; *p*-value: .012).

These data identify significant variables in relation to COVID-19 incidence rate and illustrate the trend of these variables as the COVID-19 incidence rate increases. To further analyze these trends, multiple linear regression models were run. For each response variable (incidence rate — IR; case fatality rate — CFR), a base model was run: a simple linear regression of the form 

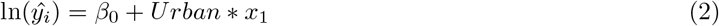

where the discrete variable *Urban* is 1 for urban counties and 0 for rural counties, in which the urban/rural status of counties is defined by the US Office of Management and Budget [21]. Table 3 presents the base, full, and rural-only incidence rate models. Three counties (Fig. 1, left map; Loving: left; Borden: center; King: right) with no confirmed cases were removed to allow for the natural log transformations. The base model regresses ln(*IR*) on *Urban*, providing a control to mathematically establish both the significance and nature of a county’s urban-rural status when *Urban* is the only predictor in consideration. Changes in the *p*-value of *Urban* as a result of adding more predictor variables to the base model highlight the importance of certain predictor variables in determining urban-rural COVID-19 risk disparities.

**Figure 1.**
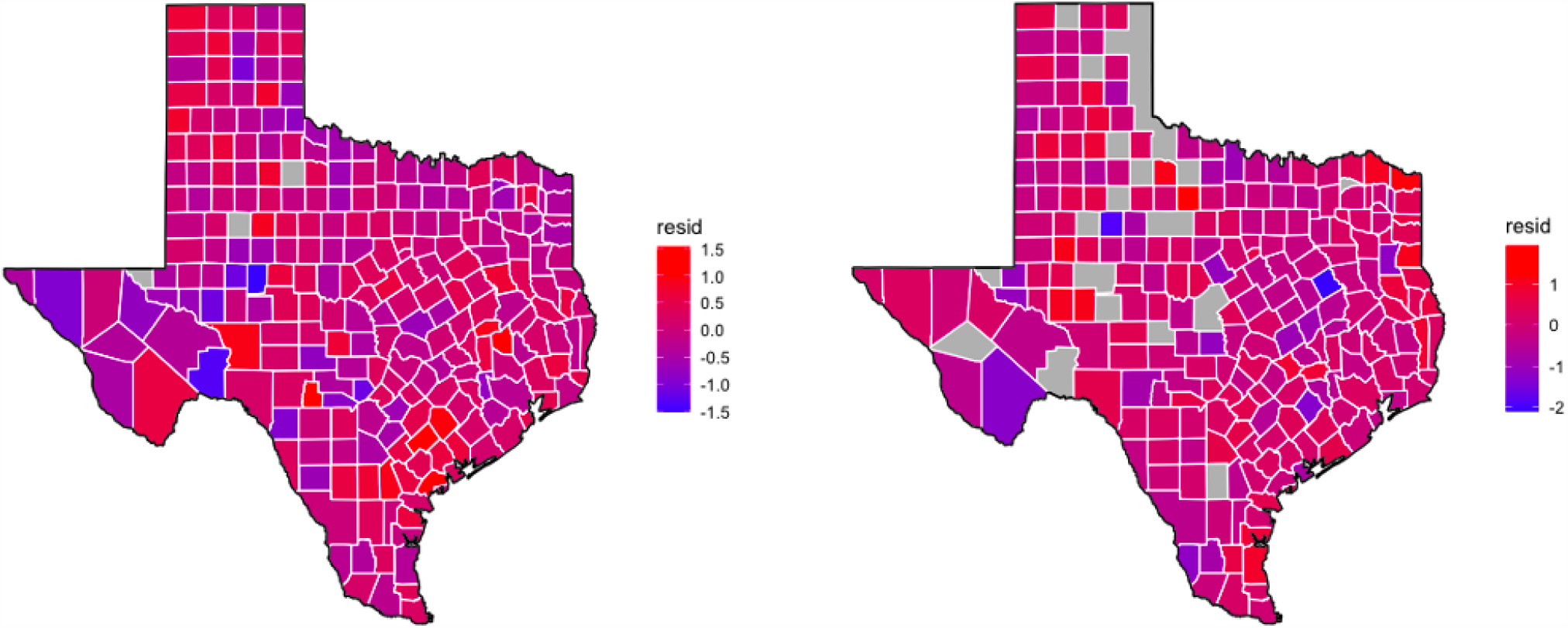
Residuals of the full incidence rate model (left) and full case fatality rate model (right). Counties with no confirmed cases are displayed in gray on the left, and counties with no deaths are displayed in gray on the right. The scales for the map on the left and the map on the right refer to ln (*IR*) and ln (*CFR*), respectively.

The IR base model predicts an incidence rate of 0.0151 in a rural county and 0.0213 in an urban county, not accounting for any other variables: a 41.1% increase. Of note, *Urban* is a highly significant variable in the base model (*p*-value: *<* .001).

According to our full incidence rate model, counties with high percentages of Hispanic and/or African American residents are expected to experience significantly higher incidence rates of COVID-19, as are counties with one or more large nursing homes (*>* 100 residents). *Hispanic* is the most significant variable in the model, with a very small *p*-value (*p*-value: *<* .001) and the largest impact on the *R*^2^ upon removal (Table 9; −19.3%). Counties with one or more large nursing homes are particularly at risk for COVID-19; specifically, a county with one or more large nursing homes (*Nursing Homes* = 1) is expected to experience a 40.6% higher incidence rate from COVID-19 than a similar county without any large nursing homes. These data underscore the importance of outbreaks in nursing homes, establishing the presence of large nursing homes as a major factor affecting the incidence rate of COVID-19 in a particular county. *Elderly* is the only predictor variable in the model with a negative coefficient, indicating that counties with a higher percentage of elderly (*>* 70 years of age) residents are expected to have lower COVID-19 incidence rates. *Black* is the variable with the largest positive coefficient; however, the standard deviation of the percentage of black residents (Table 7; 0.066) is lower than the standard deviation of Hispanic residents (Table 7; 0.233), so that *Black* has a lesser impact on ln 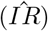 than *Hispanic*.

The objective of these MLR models is to analyze urban-rural differences, hence the presence of the variable *Urban* in both the base and full models. The *p*-value of *Urban* was significant in the base model and insignificant in the full model; adding the variables *Elderly, Nursing Homes, Hispanic* and *Black* increased the *p*-value of *Urban* from *<* .001 to 0.957, implying that the increased incidence rate of COVID-19 in urban counties is largely attributable to differences in the percentages of elderly, Hispanic, and black residents, and by the presence of large nursing homes. Adding the four aforementioned variables also hugely improved the adjusted *R*^2^ (0.052 to 0.448) and *F* statistic (14.59 to 41.61).

Full models were filtered to include only rural counties to observe any unusual shifts in coefficients or *p*-values of the variables in the full model. As expected, little difference was observed between the full and rural IR models. *Elderly* is slightly less significant in the rural model (.020 vs. .046) and *Black* has a considerably larger coefficient (2.553 vs. 3.261). No variables changed signs and there were no major changes in significance, as is consistent with the insignificance of *Urban* as a predictor variable in the full model (*p*-value: .957; Table 9; −.001%).

### Case Fatality Rate Analysis

Table 4 presents the distribution of county-level demographics and health outcomes by quintiles of COVID-19 case fatality rate in Texas on September 15, 2020. The median percentages of obese residents, unemployed residents, and diabetic residents follow generally positive trends, with medians increasing as they approach the third quintile of case fatality rate (1.9% − 2.9%) and deviating as they approach the fourth and fifth quintiles (*>* 2.9%). The percentage of elderly residents follows a strictly positive trend from the second quintile to the fifth quintile, with the first quintile deviating from this pattern. The fifth quintile of case fatality rate (*>* 4.2%) has the highest median percentage of elderly residents (median: 14.3%; *p*-value: *<* .001), lowest median percentage of Hispanic residents (median: 23.5%; *p*-value: .079), and lowest median percentage of residents ages 20-39 (median: 21.2%; *p*-value: *<* .001), in contrast with the low median percentage of elderly residents (median: 9.3%; *p*-value: *<* .001), high median percentage of Hispanic residents (median: 50.2; *p*-value: *<* .001), and high median percentage of residents ages 20-39 (median: 27.2; *p*-value: *<* .001) in the fifth quintile of COVID-19 incidence rate.

Table 5 presents the base, full, and rural-only CFR models. 28 counties with no deaths as of September 15, 2020 were not included in the CFR models to allow for a natural log transformation on the response variable. The immediate takeaway from the CFR base model is that rural counties experience higher case fatality rates from COVID-19 as opposed to lower incidence rates from COVID-19. The base model predicts a case fatality rate of 1.94% in a urban county and 2.97% in an rural county: a 53.1% increase. Once again, *Urban* is highly significant in the base model (*p*-value: *<* .001).

From our full CFR model, counties with a high percentage of elderly residents are predicted to experience a higher case fatality rate from COVID-19, as are counties with a high percentage of obese residents. Counties with a high percentage of residents ages 20-39 and, interestingly, counties with a high percentage of smokers are predicted to have lower case fatality rates. The coefficient of *Nursing Homes* is negative (−0.210), indicating that a county with one or more large nursing homes is expected to experience a 18.9% lower case fatality rate from COVID-19 than a similar county without any large nursing homes.

To provide a basis for direct comparison between case fatality rate and incidence rate of COVID-19, a model was created that regressed ln 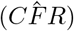 on the predictor variables from the full incidence rate model (*Urban, Elderly, Nursing Homes, Hispanic, Black*; Table 6).

The following changes were observed: 1) The coefficient of *Elderly* reversed signs, increased considerably in magnitude (IR: −2.324 to CFR: 8.079), and had a much larger impact on the *R*^2^ upon removal (Table 9; IR: −1.21% vs. CFR: −11.7%), 2) The coefficient of *Nursing Homes* reversed signs (IR: 0.343 vs. CFR: −0.251), 3) *Hispanic* became a much less significant variable (Table 9; IR: −19.3% vs. CFR: −3.49%), and 4) *Black* was not significant in the case fatality rate model (*p*-value: .527). Of note, the coefficient of *Urban* remained high (IR: 0.957 vs. CFR: 0.945), indicating that both the difference in incidence rate and the difference in case fatality rate between urban and rural counties is largely attributable to these four variables.

To visualize model performance and identify region-specific variations in prediction, we created residual maps. Fig. 1 presents the residuals of the 251 counties in the full incidence rate model (left) and the 226 counties in the full case fatality rate model (right). Of note, the full IR model generally overestimates the incidence rate of COVID-19 in the urbanized regions of the Coastal Plains, while the full CFR model underestimates the case fatality rate in these same regions. The opposite is true for counties in the Mountains and Basins region, for which the full IR model underestimates the incidence rate and the full CFR model overestimates the case fatality rate.

The full CFR model was filtered to include only rural counties to observe any unexpected differences. Similar to the rural-only incidence rate model, no variables changed signs or shifted considerably in magnitude, but *Elderly* (*p*-value: .069) and *Nursing Homes* (*p*-value: .058) were no longer significant when including only rural counties in the CFR model.

## Discussion

Our study highlights a concerning disparity between urban and rural counties, consistent with several other studies, wherein urban counties experience higher incidence rates, yet rural counties experience higher case fatality rates [2, 22]. Moreover, our study is the first to identify Hispanic & African American populations, elderly populations, and large nursing homes as major contributing factors to this urban-rural disparity in Texas. Knowing these important determinants of county health and their quantitative effect on COVID-19 incidence rate and case fatality rate makes it possible to predict which counties are at the highest risk for serious effects from COVID-19. As the counties with the highest levels of variables that positively impact the predicted COVID-19 case fatality rate are also those with lesser health resources and higher prevalence of health-related COVID-19 risk factors, it is imperative that traditionally vulnerable populations in such counties, including the elderly and immunocompromised, receive adequate essential resources from local providers and health agencies. Furthermore, the alarming positive effect of elderly populations on case fatality rate stresses the need to insulate and closely monitor counties with high percentages of elderly people. Equally concerning is the observation that rural counties, which are predicted to experience higher case fatality rates due to their high percentage of elderly residents and obese residents, lack the health resources and infrastructure of urban counties, exacerbating these effects. Streamlining hospital transportation, providing at-home medical services, and stockpiling resources for rural hospitals can lessen COVID-19 mortality in rural regions. Urban counties face a different problem, having higher incidence rates and lower case fatality rates. While urban counties have more widely available healthcare services, they also have denser populations. These denser populations, coupled with higher predicted incidence rates, have the potential to strain hospital capacities, as demonstrated in mid-July. High incidence rates in urban counties are linked to lower percentages of elderly residents, higher percentages of black residents, and presence of large nursing homes. Of these, the variable that can most reasonably be addressed is nursing homes. The significance of COVID-19 outbreaks in nursing homes is widely known, and such outbreaks are often a result of a lack of testing and close contact with healthcare providers. A plausible measure could be to conduct consistent facility-wide testing, which has been shown to prevent outbreaks in nursing homes [23].

In addition to analyzing COVID-19 incidence rate and case fatality rate, our study uncovered interesting behavior in the predictor variables analyzed. *Black* is not significant in the case fatality rate model and has a very low *R*^2^ upon removal, in contrast with its significance in the incidence rate model. This indicates that while counties with a high percentage of black residents have higher incidence rates, they do not necessarily have higher case fatality rates, despite these counties having higher rates of poverty and lesser access to healthcare. Counties with many Hispanic residents experience both higher incidence rates and higher case fatality rates, which can be partially attributed to high levels of obesity and diabetes coupled with low rates of health insurance coverage and higher representation in “superspreader” sites such as prisons or homeless shelters [24, 25]. Furthermore, Hispanic residents face language barriers and immigration status, which can complicate the process of obtaining medical aid [26, 27]. This highlights the concerning vulnerability of Hispanic populations along with the importance of adequate communication and provision of medical aid during this time. The negative coefficient of *Smoking* in the case fatality rate model was unexpected, as smoking is a CDC-defined risk factor for COVID-19 and prevalence of smoking in a county is positively correlated with elderly populations and with the incidence of respiratory disease, hypertension, obesity, and cardiovascular disease in Texas. Our study is not the first to report this relationship: Norden et al. [28] at Stanford University and the University of Washington found a similar negative relationship between smoking and COVID-19 case fatality rate across multiple countries, controlling for sex ratio, obesity, temperature, and elderly populations. In their study, several biological explanations for a protective effect of smoking on COVID-19 fatality were proposed. While the possibility of a true protective effect of smoking on COVID-19 can not be ruled out, it is likely that a confounding variable, a variable not accounted for in our model, the effects of county-level data aggregation, or an interaction between another predictor variable and smoking is responsible for the negative coefficient. Regardless, the magnitude of the negative effect of smoking on COVID-19 case fatality rate prompts further exploration to clarify the role of smoking in COVID-19 fatality, both biologically and statistically.

The variables included in our full models are those that are the statistically strongest in predicting COVID-19 risk. In creating our models, we were surprised to find that health access (health insurance, primary care physicians), financial status (median income, poverty levels), mortality rate from respiratory disease, and population density were not among the best predictors, as these variables are thought to have significant effects on COVID-19 risk. However, several variables (*Elderly, Hispanic, Nursing Homes*) made consistent appearances, decreasing the likelihood that these variables were merely acting as proxies for more significant variables and indicating that there is some intrinsic aspect of populations of elderly/Hispanic residents, apart from correlations with other variables, that affects COVID-19 risk. This emphasizes the need to ensure that these populations are properly insulated against COVID-19 outbreaks and establishes them as important factors in population heterogeneity in relation to COVID-19 risk. Moreover, the residuals of our models contain important information about which counties are performing better or worse than expected. Comparing counties with negative residuals for common preventative measures could reveal which prevention strategies are most effective, which could then be implemented in counties with high residuals.

There exist a number of limitations in our study and opportunities for further work. Several counties with a value of 0 for the response variable were removed from our models to allow for a log transformation on the response variable. Excluding counties with no cases or deaths causes our study to underestimate the impact of variables that may be particularly significant in those counties, potentially leading to the omission of important variables in determining incidence rate or case fatality rate from COVID-19 and lowering the accuracy of prediction in counties with no deaths and/or confirmed cases. As the pandemic progresses, this limitation may fade. Another limitation was the designation of urban/rural and variables for super spreading contexts as discrete variables. In reality, the urban/rural designation is not black or white. Many counties cannot be classified as definitively urban or rural, with some sections being urban and others rural. A simple discrete variable does not capture this variability, and a more proper designation may be large metropolitan/midsize metro/small metro/micropolitan/semirural/rural. Regarding super spreading contexts, a discrete variable does not adequately convey the extent to which the specific super spreading context impacts the county, and a variable that captures the proportion of people in the population who are employed or reside in a super spreading context may be a more robust predictor. Future research can extend the premise of our models to investigate more response variables, including *R*_*t*_, testing rates, and possibly hospital capacity. By identifying at-risk counties and predicting the relative magnitude of county-level COVID-19 incidence rate or case fatality rate based on quantifiable population characteristics, resources can be more appropriately allocated to decrease the disproportionate impact of COVID-19 in Texas and a groundwork can be laid for future population studies to explore the relationships we discovered in depth.

## Data Availability

The codes and data used to generate our results are available from the corresponding authors upon reasonable request.

## Author contributions

AW designed the study. AKL, SZ, CS, and JW were responsible for data curation and model creation. AKL conducted statistical analysis and produced tables and figures. All authors contributed towards interpretation of study findings. AKL wrote the first draft, and all authors critically reviewed and approved the final contents of the paper.

## Acknowledgements

A big thanks to Texas State University and the Mathworks Honors Summer Math Camp for providing a platform for us to collaborate virtually to undertake this research. We’d also like to thank Dr. Lauren Ancel-Meyers and the UT COVID-19 Modeling Consortium for sharing their knowledge and advice on a daily basis, and for providing inspiration and data leads for this project.

## Appendices

**Table 1.**
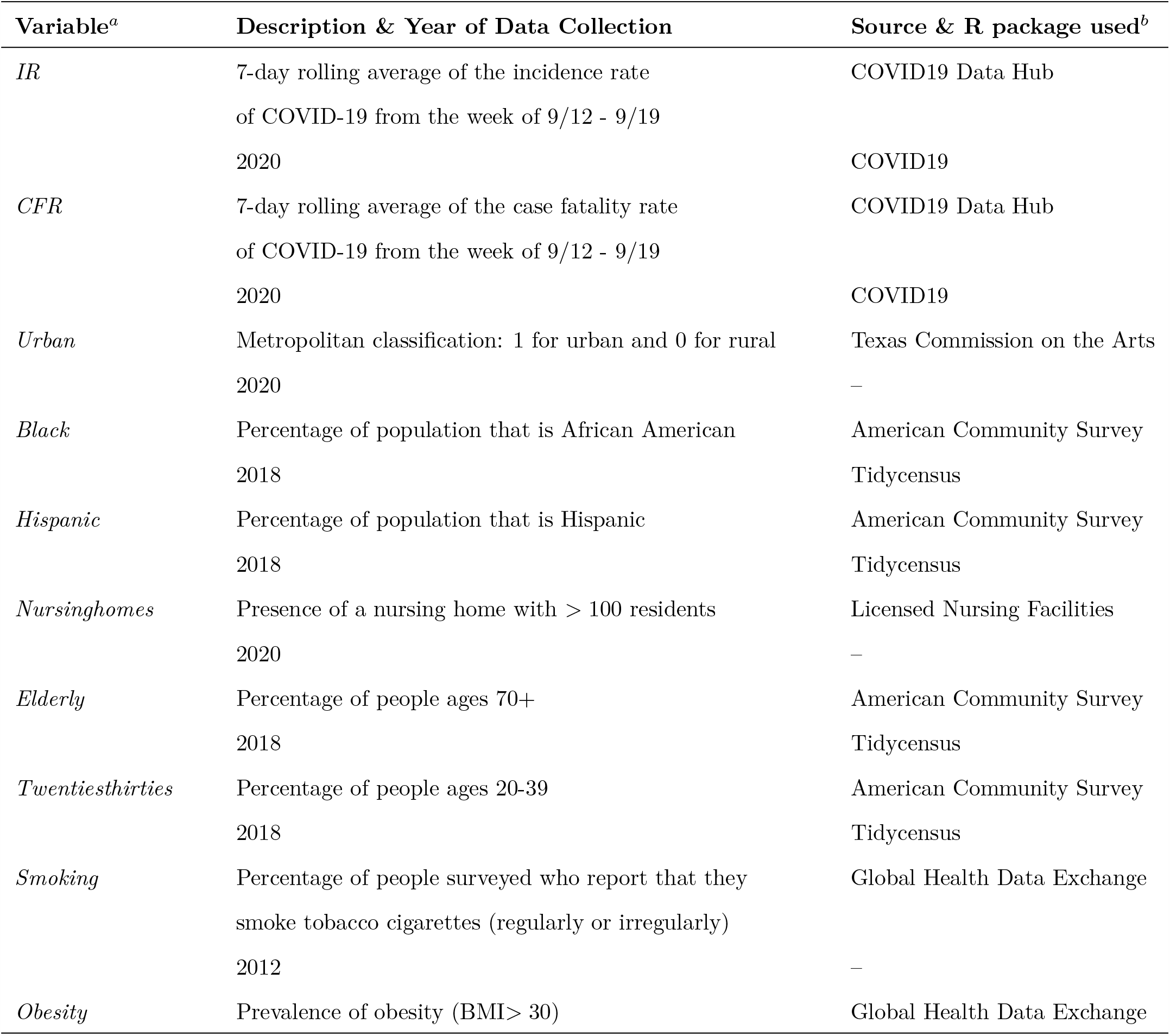
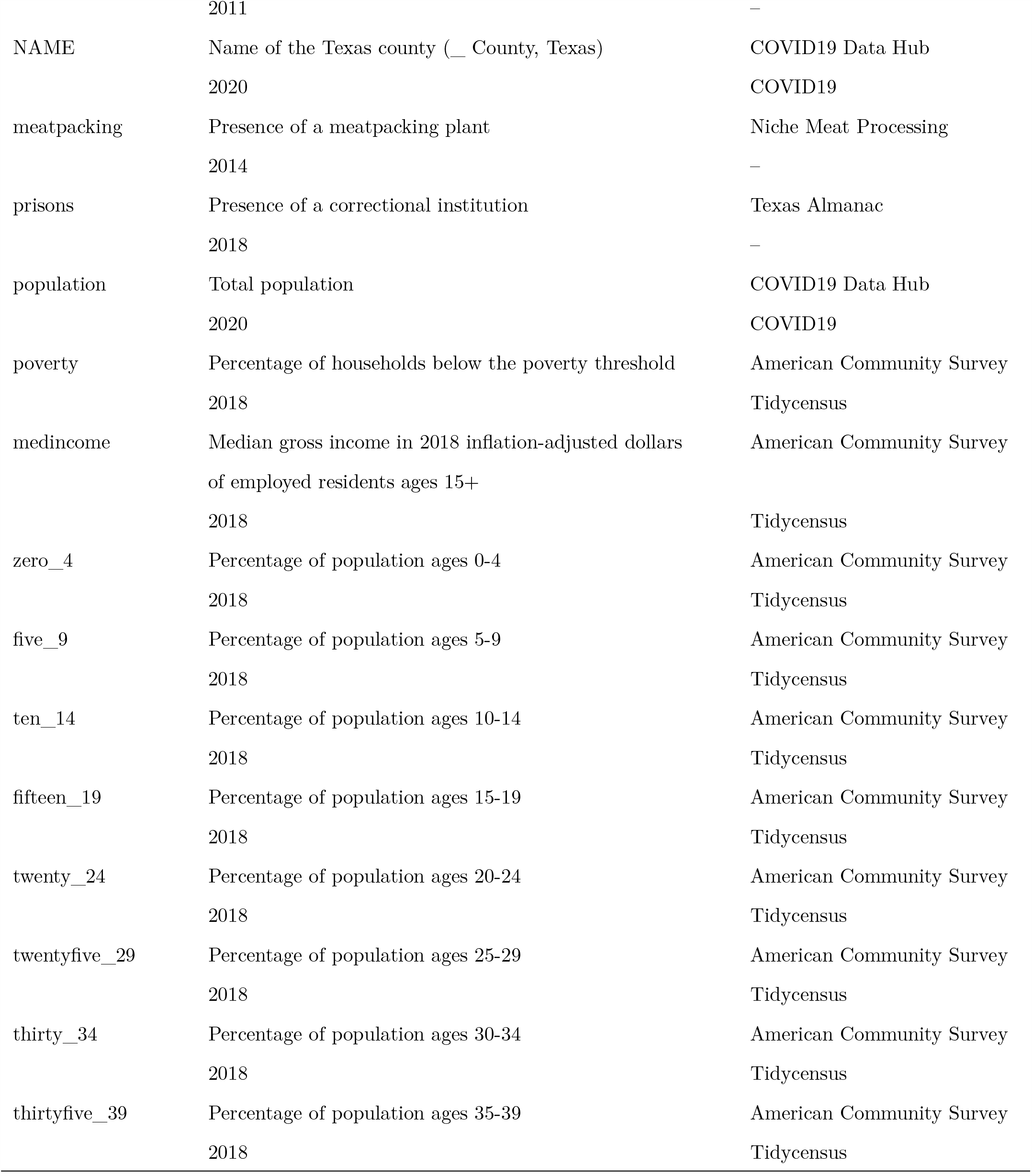
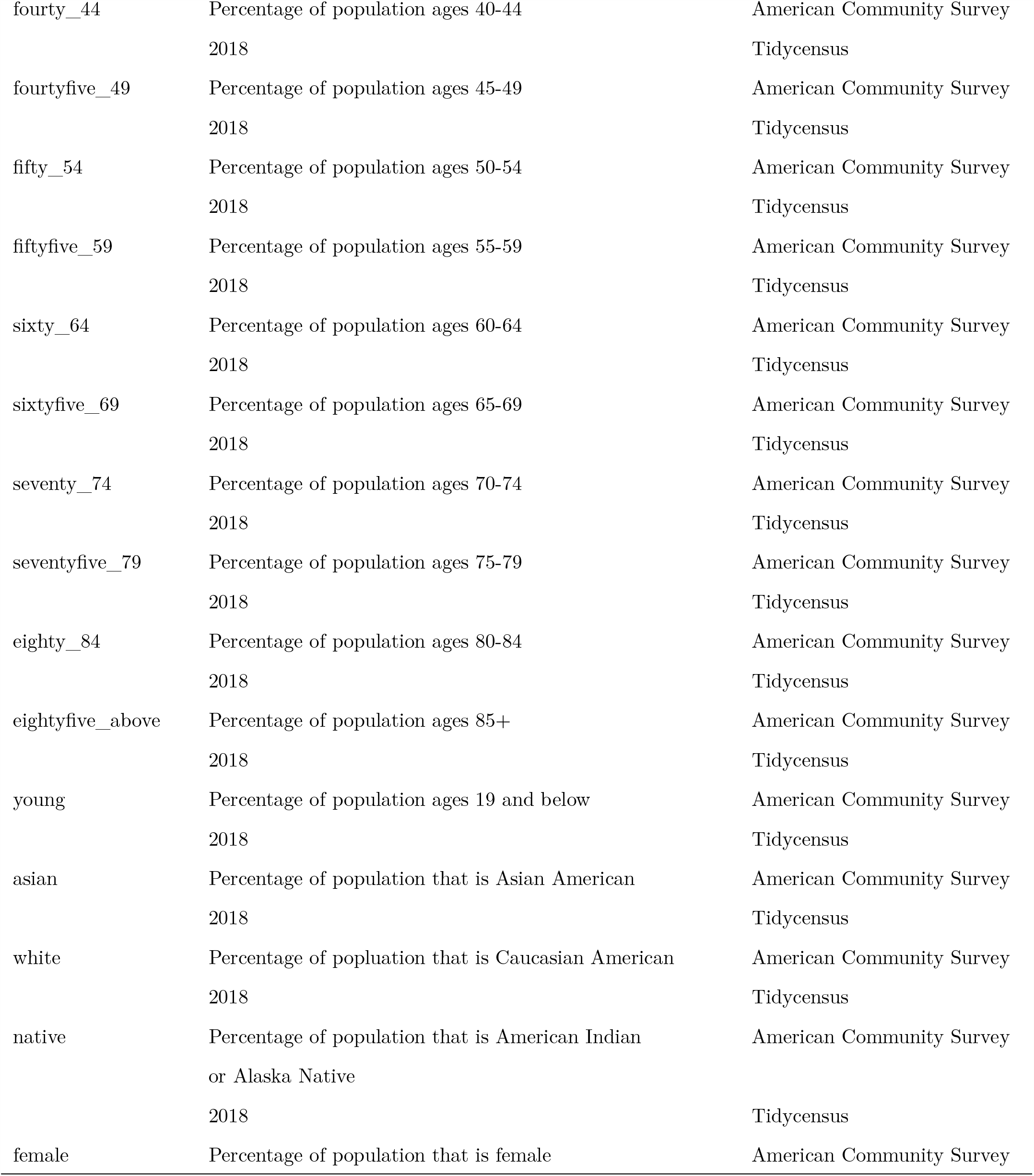
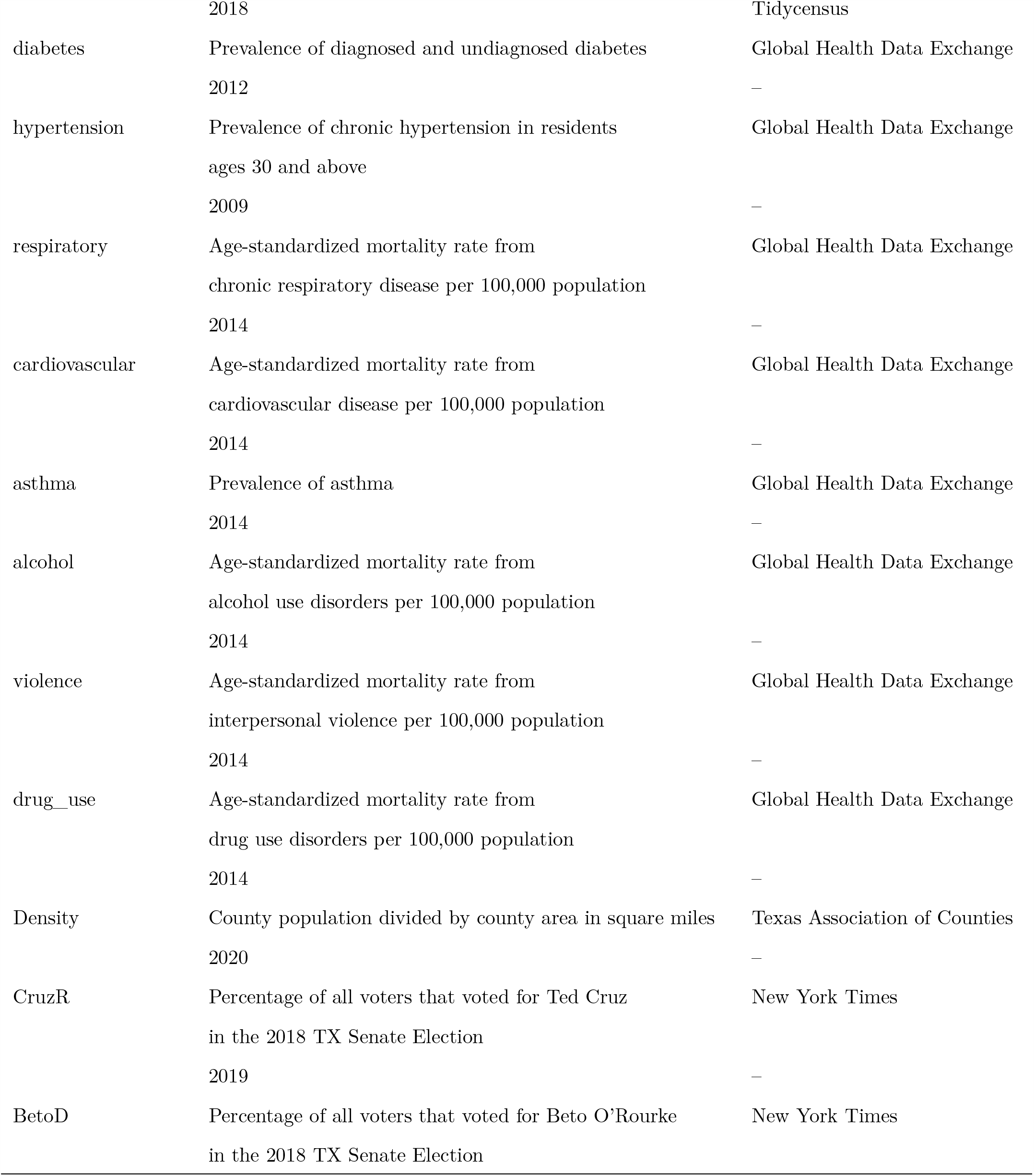
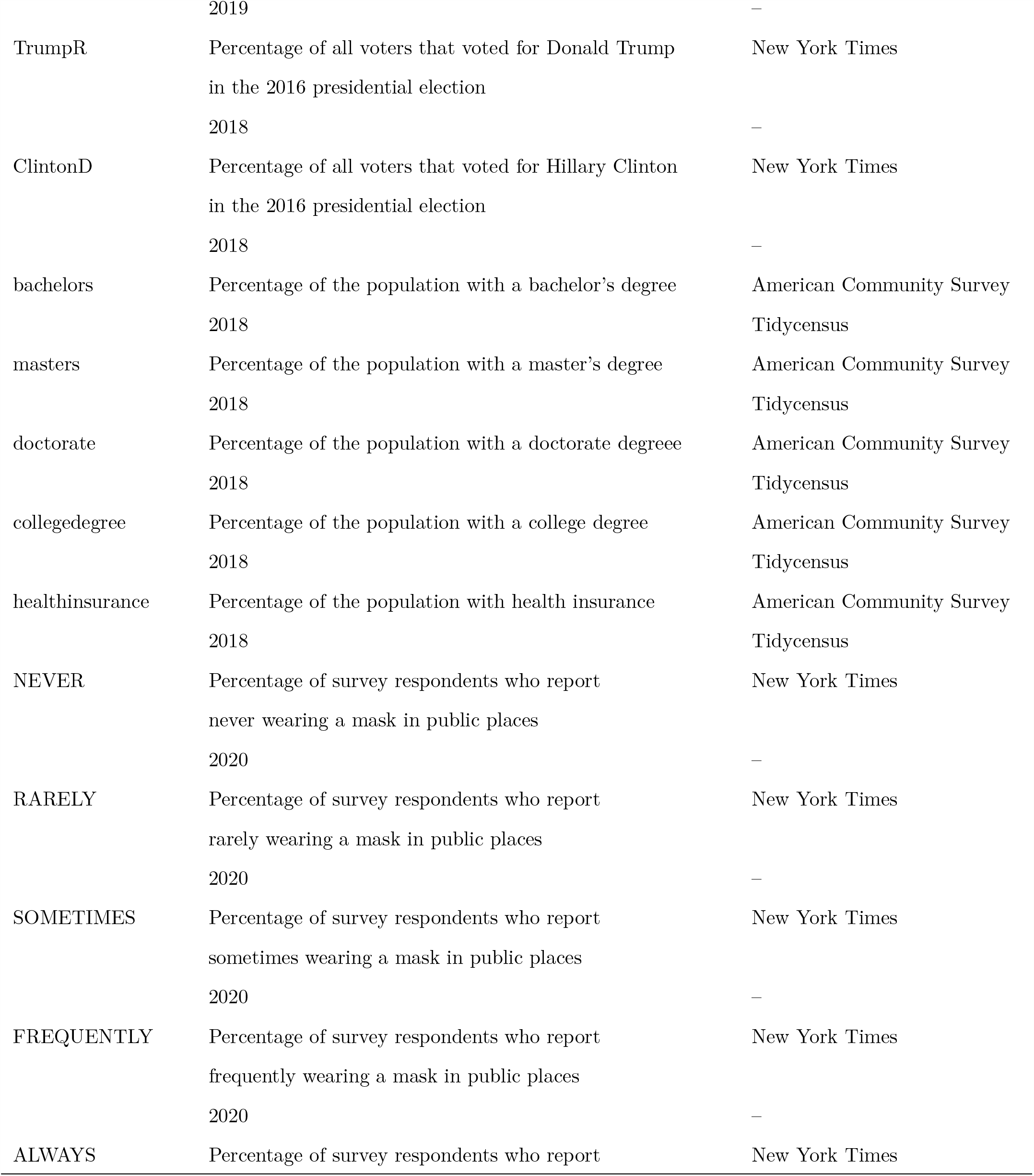
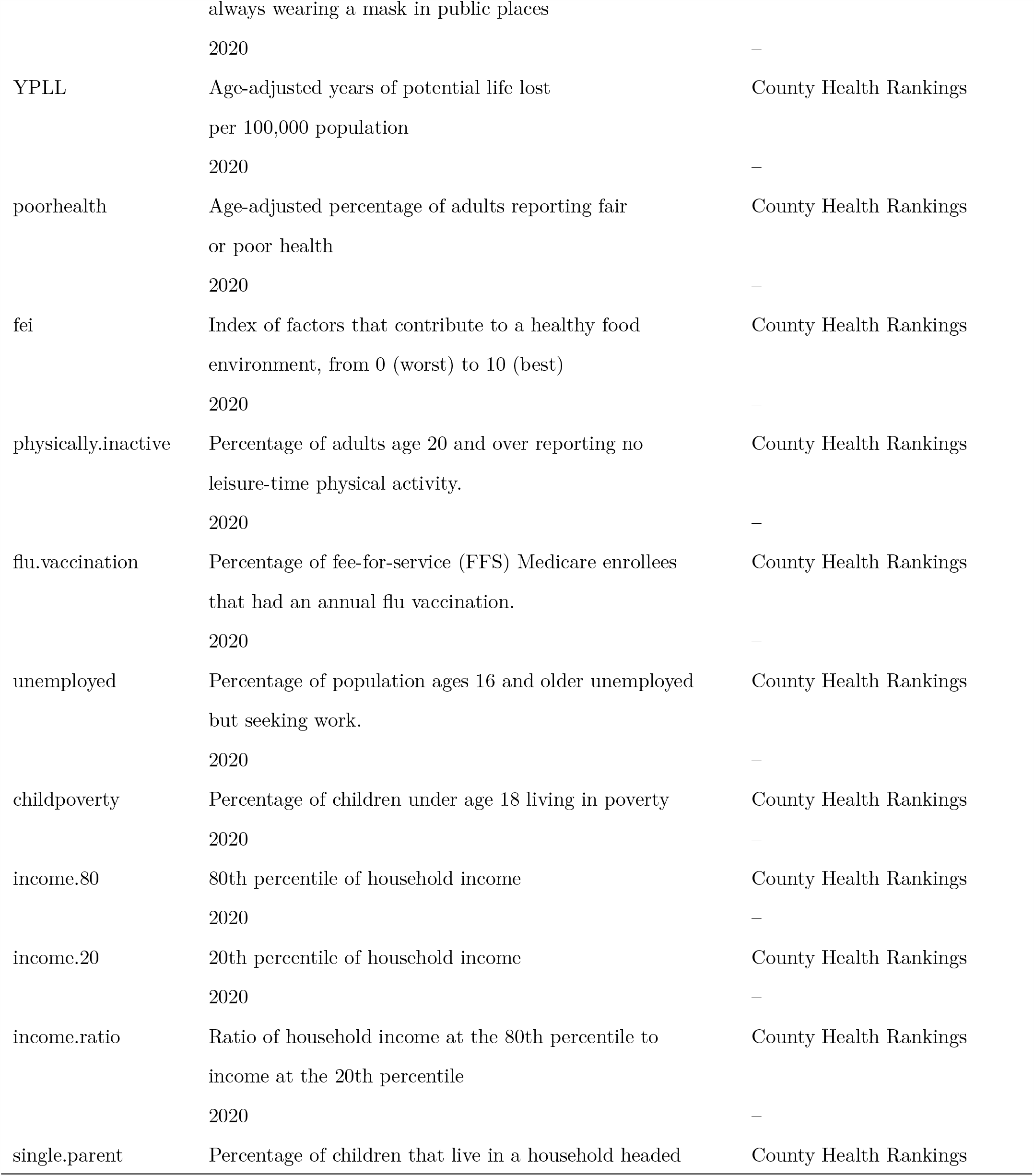
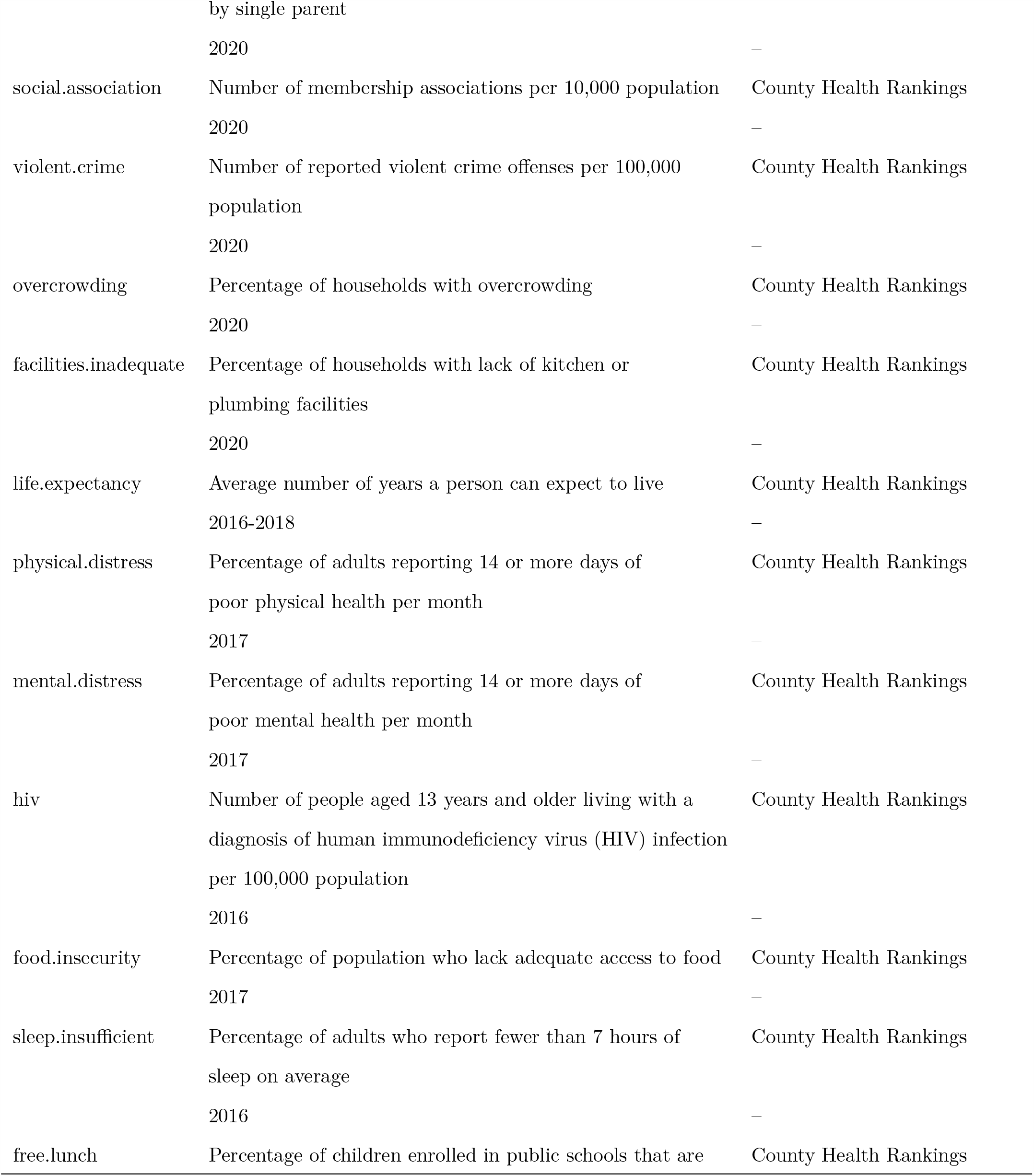
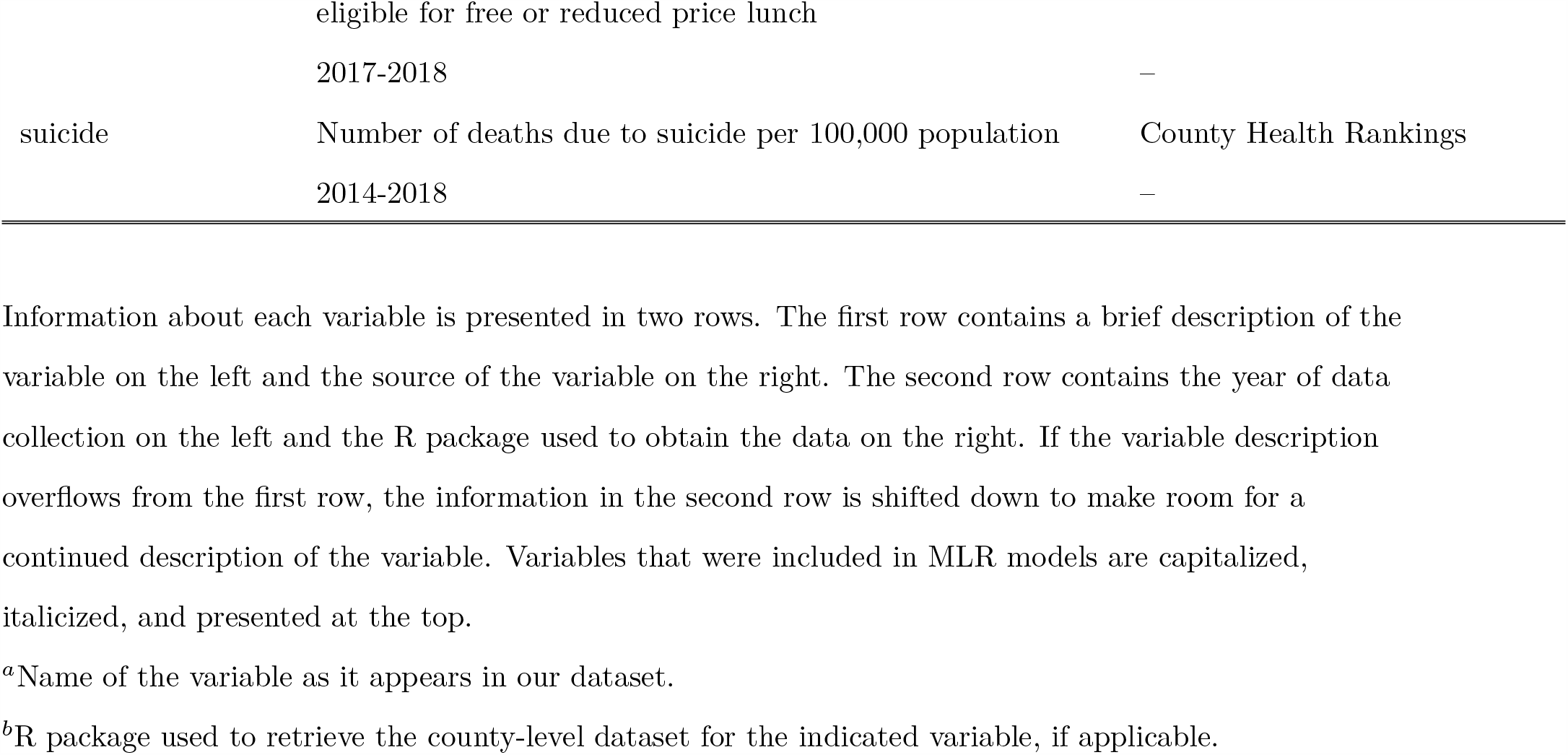
Data Descriptions and Sources

**Table 2.** Texas County-Level Variable Comparisons by Quintiles of COVID-19 Incidence Rate through September 15, 2020^a^

| Incidence Rate | 0.00-964.7 | 964.7-1426.7 | 1426.7-1923.6 | 1923.6-2816.3 | >2816.3 | *p*-value<sup>b</sup> |
| --- | --- | --- | --- | --- | --- | --- |
| Data presented as median (IQR) |  |  |  |  |  |  |
| Counties | 51 | 51 | 50 | 51 | 51 | – |
| *Demographics* |  |  |  |  |  |  |
| Black | 1.8 (3.8) | 4.6 (8.0) | 4.9 (10.3) | 6.3 (8.7) | 4.8 (8.6) | 0.002 |
| White | 90.2 (9.2) | 86.7 (12.1) | 85.8 (11.5) | 81.8 (15.1) | 83.4 (14.8) | 0.002 |
| Hispanic | 18.5 (18.3) | 19.3 (24.6) | 23.5 (17.2) | 36.8 (33.9) | 50.2 (38.6) | <.001 |
| Elderly | 14.4 (5.6) | 12.7 (5.9) | 12.5 (4.7) | 9.8 (4.1) | 9.3 (2.3) | <.001 |
| Age 20-39 | 20.8 (4.7) | 23.1 (6.1) | 23.1 (4.2) | 25.9 (5.2) | 27.2 (5.6) | <.001 |
| Female | 49.9 (3.3) | 50.3 (1.8) | 50.4 (2.0) | 49.7 (2.0) | 49.4 (3.3) | 0.097 |
| *Health Status* |  |  |  |  |  |  |
| Obesity | 37.6 (3.1) | 38.9 (3.5) | 39.6 (3.5) | 39.3 (3.3) | 40.3 (3.3) | <.001 |
| Smoking | 21.2 (4.4) | 22.4 (3.7) | 21.4 (4.3) | 20.8 (5.4) | 20.6 (3.2) | 0.108 |
| Diabetes | 14.4 (1.2) | 15.6 (2.2) | 15.4 (2.2) | 16.2 (2.2) | 17.0 (3.8) | <.001 |
| Health Insurance | 98.8 (1.8) | 98.7 (1.7) | 98.8 (1.2) | 98.8 (2.1) | 97.7 (6.2) | 0.012 |
| *Living Conditions* |  |  |  |  |  |  |
| Unemployed | 3.4 (0.6) | 3.5 (1.0) | 3.7 (0.9) | 3.6 (1.4) | 3.8 (1.7) | 0.078 |
| Poverty | 13.5 (5.8) | 14.1 (5.5) | 15 (5.0) | 14.4 (6.4) | 16.6 (4.9) | 0.002 |
| Density | 5.5 (19.7) | 27.4 (63.3) | 19.5 (42.2) | 40.1 (169.4) | 36.2 (63.7) | <.001 |

<sup>a</sup>County-level incidence rate calculated as the 7-day rolling average of confirmed COVID-19 cases divided by population. Expressed per 100,000 population on September 15, 2020.
<sup>b</sup>p-values determined using the Kruskal-Wallis test. p-value < 0.05 is considered significant.

**Table 3.**
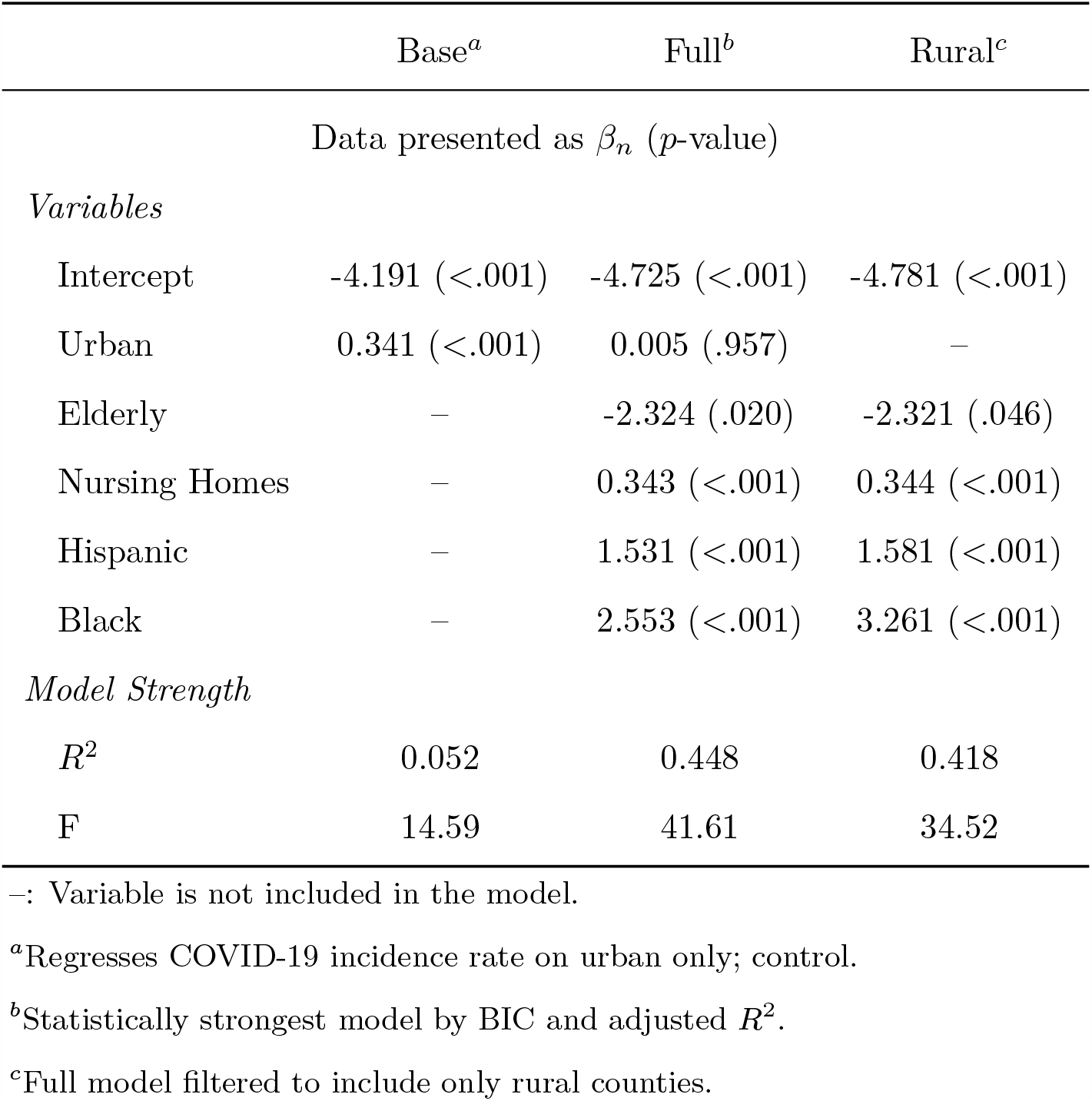
COVID-19 Incidence Rate Models

**Table 4.** Texas County-Level Variable Comparisons by Quintiles of COVID-19 Case-Fatality Rate through September 15, 2020^a^

| Case Fatality Rate | 0%-1.3% | 1.3%-1.9% | 1.9%-2.9% | 2.9%-4.2% | >4.2% | p-value<sup>b</sup> |
| --- | --- | --- | --- | --- | --- | --- |
| Data presented as median (IQR) |  |  |  |  |  |  |
| Counties | 50 | 50 | 50 | 50 | 51 | — |
| *Demographics* |  |  |  |  |  |  |
| Black | 3.8 (8.6) | 7.2 (10.4) | 5.3 (5.8) | 2.5 (5.8) | 3.0 (6.8) | 0.004 |
| White | 85.6 (10.3) | 81.7 (16.2) | 87 (10.4) | 88.3 (12.1) | 88.0 (13.3) | 0.142 |
| Hispanic | 26.9 (20.3) | 23.9 (9.8) | 40.3 (39.7) | 39.3 (39.5) | 23.5 (40.9) | 0.079 |
| Elderly | 11.3 (6.4) | 9.9 (4.4) | 10.5 (4.4) | 11.4 (3.3) | 14.3 (6.1) | < .001 |
| Age 20-39 | 24.6 (9.0) | 25.8 (5.2) | 25.2 (5.7) | 24.6 (4.1) | 21.2 (5.5) | < .001 |
| Female | 49.4 (4.0) | 50.3 (1.9) | 50.2 (2.2) | 50.4 (2.9) | 50.4 (1.9) | 0.155 |
| *Health Status* |  |  |  |  |  |  |
| Obesity | 37.7 (3.3) | 38.5 (3.8) | 39.9 (2.3) | 39.6 (2.7) | 39.7 (3.1) | < .001 |
| Smoking | 20.9 (3.5) | 21.3 (3.5) | 22.1 (4.2) | 21.5 (5.1) | 20.7 (3.7) | 0.455 |
| Diabetes | 15.0 (1.8) | 15.1 (2.4) | 16.4 (2.3) | 16.1 (3.1) | 16.1 (2.6) | < .001 |
| Health Insurance | 98.5 (6.8) | 98.8 (3.3) | 98.8 (1.2) | 98.4 (3.9) | 98.6 (1.8) | 0.645 |
| *Living Conditions* |  |  |  |  |  |  |
| Unemployed | 3.3 (0.7) | 3.4 (1.0) | 3.7 (1.2) | 3.7 (1.3) | 4.1 (1.8) | < .001 |
| Poverty | 12.5 (4.6) | 13.5 (5.3) | 15.5 (3.5) | 15.8 (5.9) | 16.8 (7.7) | < .001 |
| Density | 9.4 (25.9) | 48.8 (170.1) | 39.5 (69) | 26.1 (43.1) | 11.5 (27.2) | < .001 |

<sup>a</sup> County-level case fatality rate calculated as the 7-day rolling average of COVID-19 deaths divided by the 7-day rolling average of confirmed COVID-19 cases.
<sup>b</sup> p-values determined using the Kruskal-Wallis test. p-value < 0.05 is considered significant.

**Table 5.** COVID-19 Case Fatality Rate Models

|  | Base<sup>a</sup> | Full<sup>b</sup> | Rural<sup>c</sup> |
| --- | --- | --- | --- |
| Data presented as $\beta_n$ ($p$-value) | | | |
| *Variables* |  |  |  |
| Intercept | -3.516 (<.001) | -4.113 (<.001) | -4.113 (<.001) |
| Urban | -0.426 (<.001) | 0.030 (.773) | — |
| Elderly | — | 3.876 (.015) | 3.545 (.069) |
| Nursing Homes | — | -0.210 (.013) | -0.186 (.058) |
| Age 20-39 | — | -4.347 (<.001) | -4.633 (.003) |
| Obesity | — | 6.354 (<.001) | 6.485 (.001) |
| Smoking | — | -4.881 (<.001) | -5.752 (.002) |
| *Model Strength* |  |  |  |
| $R^2$ | 0.086 | 0.332 | 0.238 |
| F | 21.06 | 19.62 | 11.10 |

—: Variable is not included in the model.
<sup>a</sup>Regresses case fatality rate on urban only; control.<sup>b</sup>Model with the lowest BIC (Bayesian Information Criteria).<sup>c</sup>Full model filtered to include only rural counties.

**Table 6.** Case Fatality Rate Model with Full Incidence Rate Model Predictors

|  | IR | CFR |
| --- | --- | --- |
| *Variables* |  |  |
| Intercept | −4.725 (<.001) | −4.698 (<.001) |
| Urban | 0.005 (.957) | 0.007 (.945) |
| Elderly | −2.324 (.020) | 8.079 (<.001) |
| Nursing Homes | 0.343 (<.001) | −0.251 (.006) |
| Hispanic | 1.531 (<.001) | 0.685 (.002) |
| Black | 2.553 (<.001) | 0.435 (.527) |
| *Model Strength* |  |  |
| $R^2$ | 0.463 | 0.235 |
| F | 43.81 | 14.79 |

*CFR* is regressed on the predictor variables from the full IR model: *Urban*, *Elderly*, *Nursing Homes*, *Hispanic*, and *Black*. Full IR model is provided for comparison.

**Table 7.** Summary Statistics for Incidence Rate Models

|  | Mean | SD<sup>a</sup> |
| --- | --- | --- |
| Incidence Rate |  |  |
| Urban | 0.024 | 0.012 |
| Rural | 0.019 | 0.012 |
| *Overall*<sup>b</sup> | 0.020 | 0.012 |
| Black |  |  |
| Urban | 0.098 | 0.077 |
| Rural | 0.053 | 0.058 |
| *Overall* | 0.064 | 0.066 |
| Hispanic |  |  |
| Urban | 0.325 | 0.234 |
| Rural | 0.359 | 0.233 |
| *Overall* | 0.351 | 0.233 |
| Elderly |  |  |
| Urban | 0.090 | 0.026 |
| Rural | 0.129 | 0.038 |
| *Overall* | 0.119 | 0.039 |
| Nursing Homes |  |  |
| Urban | 0.968 | 0.177 |
| Rural | 0.436 | 0.498 |
| *Overall* | 0.570 | 0.496 |

<sup>a</sup>Standard deviation.
<sup>b</sup>Includes $N = 251$ counties in the full IR model.

**Table 8.** Summary Statistics for Case Fatality Rate Models

|  | Mean | SD<sup>a</sup> |  | Mean | SD |
| --- | --- | --- | --- | --- | --- |
| CFR |  |  | Black |  |  |
| Urban | 0.022 | 0.011 |  | 0.098 | 0.076 |
| Rural | 0.038 | 0.033 |  | 0.057 | 0.060 |
| *Overall<sup>b</sup>* | 0.033 | 0.029 |  | 0.068 | 0.067 |
| Hispanic |  |  | Nursing Homes |  |  |
| Urban | 0.325 | 0.234 |  | 0.968 | 0.177 |
| Rural | 0.372 | 0.242 |  | 0.491 | 0.501 |
| *Overall* | 0.359 | 0.240 |  | 0.624 | 0.485 |
| Elderly |  |  | Age 20-39 |  |  |
| Urban | 0.090 | 0.026 |  | 0.276 | 0.041 |
| Rural | 0.127 | 0.036 |  | 0.238 | 0.045 |
| *Overall* | 0.116 | 0.037 |  | 0.249 | 0.047 |
| Obesity |  |  | Smoking |  |  |
| Urban | 0.382 | 0.037 |  | 0.203 | 0.034 |
| Rural | 0.399 | 0.026 |  | 0.217 | 0.028 |
| *Overall* | 0.394 | 0.030 |  | 0.213 | 0.030 |

<sup>a</sup>Standard deviation.<sup>b</sup>Includes $N = 226$ counties in the full CFR model.

**Table 9.** Impact of Each Predictor Variable on the R^2^ Upon Removal

|  | Incidence Rate | CFR - IR Predictors | CFR - Full |
| --- | --- | --- | --- |
| Urban | −.001% | −.003% | −.024% |
| Elderly | −1.21% | −11.7% | −1.79% |
| Hispanic | −19.3% | −3.49% | — |
| Nursing Homes | −5.25% | −2.70% | −1.85% |
| Black | −4.85% | −.142% | — |
| Smoking | — | — | −4.18% |
| Obesity | — | — | −7.23% |
| Age 20-39 | — | — | −4.20% |

Each cell is the negative impact that the respective variable has on the $R^2$ of the indicated model upon removal. Percentages are differences from the $R^2$ , and are not percentages of the $R^2$ .

